# The Aging Epigenome: Integrative Analyses Reveal Functional Overlap with Alzheimer’s Disease

**DOI:** 10.1101/2025.06.08.25329218

**Authors:** Wei Zhang, David Lukacsovich, Juan I. Young, Lissette Gomez, Michael A. Schmidt, Brian W. Kunkle, Xi Chen, Eden R. Martin, Lily Wang

## Abstract

Aging is the strongest risk factor for Alzheimer’s disease (AD), yet the role of age-associated DNA methylation (DNAm) changes in blood and their relevance to AD remains poorly understood. In this study, we performed a meta-analysis of blood DNAm samples from 475 dementia-free subjects aged over 65 years across two independent cohorts, the Framingham Heart Study (FHS) at Exam 9 and the Alzheimer’s Disease Neuroimaging Initiative (ADNI). After adjusting for age, sex, and immune cell type proportions, and correcting for batch effects and genomic inflation, we identified 3758 CpGs and 556 differentially methylated regions (DMRs) consistently associated with aging in both cohorts at a 5% false discovery rate. Our pathway enrichment analyses highlighted immune response, metabolic regulation, and synaptic plasticity, all of which are key biological processes implicated in AD. Moreover, our colocalization analysis revealed 32 genomic regions where shared genetic variants influenced both DNAm and dementia risk. Adjusting for age and other covariate variables, we found roughly one-third of aging-associated CpGs are also associated with AD or AD neuropathology in independent studies external to the ADNI and FHS datasets. Finally, we prioritized 9 aging-associated CpGs, located in promoter regions of *PDE1B, ELOVL2, PODXL2*, and other genomic regions, that showed strong positive blood-to-brain methylation concordance, as well as association with AD or AD neuropathology in independent studies, after adjusting for age and other covariates. Our findings provided insights into the functional overlap between the aging processes and AD, and nominated promising blood-based biomarkers for future AD research.

## INTRODUCTION

Alzheimer’s disease (AD) is the leading cause of dementia worldwide, it already affects more than 6 million people in the United States alone; its prevalence is projected to grow to 13.8 million by 2060 as the population ages.^1^ Chronological age remains the single strongest non-modifiable risk factor, with the majority of cases occurring after age 65. Understanding the molecular changes that accompany “normal” aging, and how they intersect with pathologic processes that culminate in AD, is therefore central to both risk stratification and early-intervention strategies.

Epigenetic modifications, particularly DNA methylation (DNAm), accumulate throughout life and influence transcriptional programs that underlie immune, metabolic, and neurodegenerative pathways. ^2^ Multi-tissue “epigenetic clocks” such as Horvath’s 353-CpG model demonstrate that a subset of age-related DNAm changes faithfully track biological aging across diverse tissues. ^3^ In addition, large-scale blood epigenome-wide association studies (EWAS) have confirmed thousands of CpG sites and differentially methylated regions (DMRs) with reproducible age-dependent shifts. ^4-7^ Growing literature also links DNAm signatures both to incident dementia^8^ and to early alterations in AD neuropathology ^9^. However, because most AD EWAS treat age as a confounder to be adjusted for, the specific contribution of age-related DNAm changes in AD remains unclear.

Several recent studies have explored the molecular intersections of aging and AD. In the temporal cortex, age-related and AD-associated transcriptional shifts show extensive concordance, with most genes changing in the same direction. ^10^ Meng et al. (2016) further demonstrated that epigenomic aging signatures across multiple brain regions converge with AD-responsive genes in immune and developmental pathways.^11^ In Li et al. (2016), genome-wide blood DNAm profiles from aging individuals were systematically compared to brain DNAm profiles of AD patients, and the authors provided the first evidence that peripheral epigenetic aging signatures partially overlap with AD-associated changes in the brain. ^12^

However, these previous studies have used DNAm samples measured by the older 27k/450k Illumina arrays, and did not directly compare blood DNAm differences in aging with blood DNAm differences in AD. To address these gaps, we performed a comprehensive analysis to examine aging-associated blood DNAm and its implications for dementia, by leveraging DNAm data measured in blood from 475 dementia-free adults older than 65 years. These data were generated from two large clinical cohorts, the Framingham Heart Study (Exam 9) and the Alzheimer’s Disease Neuroimaging Initiative (ADNI). All the DNAm samples were measured using the Illumina HumanMethylation EPIC beadchip, which included more than 850,000 CpGs^13^.

We performed a meta-analysis to identify age-associated CpGs and DMRs, mapped their genomic distributions, and evaluated their transcriptional and functional relevance by integrative eQTm (DNAm-to-gene expression associations), brain-to-blood correlations in DNA methylation levels, and pathway analyses. In addition, we explored shared genetic architecture of the aging processes and dementia, by integrating our results with large-scale mQTL resources and genome-wide association summary statistics from recent dementia GWAS, and performing colocalization analysis. Finally, we compared our findings with independent AD methylation studies external to FHS and ADNI datasets.

## RESULTS

### Study datasets

Our meta-analysis included 475 participants from two cohorts: 282 individuals from the FHS Offspring cohort at Exam 9 (FHS9) and 193 individuals from the ADNI. In ADNI, we analyzed each participant’s earliest visit with available DNAm data. In FHS9, to avoid inflation due to family structure, we selected only one individual per family, prioritizing the sample with the highest bisulfite conversion rate.

The average follow-up durations for the subjects were 5.0 ± 2.3 years in FHS9 and 5.9 ± 3.0 years in ADNI. We excluded individuals who developed dementia during the follow-up period. All participants were over 65 years of age. In FHS9, the average age was 74.3 ± 6.6 years, with 56.7% females. In ADNI, the average age was 77.0 ± 6.5 years, with 50.8% females (Table 1).

**Table 1.** Characteristics of subjects included in the meta-analysis of the Framingham Heart Study Exam 9 (FHS9) and Alzheimer’s Disease Neuroimaging Initiative (ADNI) cohorts.

| Characteristics | FHS9<br>(n =282) | ADNI<br>(n = 193) |
| --- | --- | --- |
| Follow up period in years |  |  |
| Av. (sd) | 4.95 (2.31) | 5.87 (3.01) |
| Age, av. (sd) | 74.29 (6.57) | 76.97 (6.50) |
| Sex, n (%) |  |  |
| Female | 160 (56.74) | 95 (50.78) |
| Male | 122 (43.26) | 98 (49.22) |

### Genomic distribution of aging-associated blood DNAm differences at individual CpGs and differentially methylated regions (DMRs)

After adjusting for age, sex, and immune cell type proportions, and correcting for batch effects and genomic inflation (see Methods), we identified 3758 CpGs with a nominal *P-*value < 1×10⁻⁵ and a false discovery rate (FDR) < 0.05 using the inverse-variance fixed-effects meta-analysis (Figure 1, Supplementary Table 1, Table 2). All these CpGs had a consistent direction of change in the FHS9 and ADNI datasets. Among them, about half (55.7%, 2092 CpGs) were hypermethylated with increasing chronological age, and more than half (64.4%, 2,422 CpGs) were located within CpG islands or shores. Notably, among the 2092 hypermethylated CpGs, the majority (64.1%, 1340 CpGs) were located in gene promoter regions (< 2 kb from the transcription start site, TSS). On the other hand, among the 1666 hypomethylated CpGs, the majority (74.2%, 1236 CpGs) were found in distal regions (> 2 kb from the TSS).

**Figure 1.**
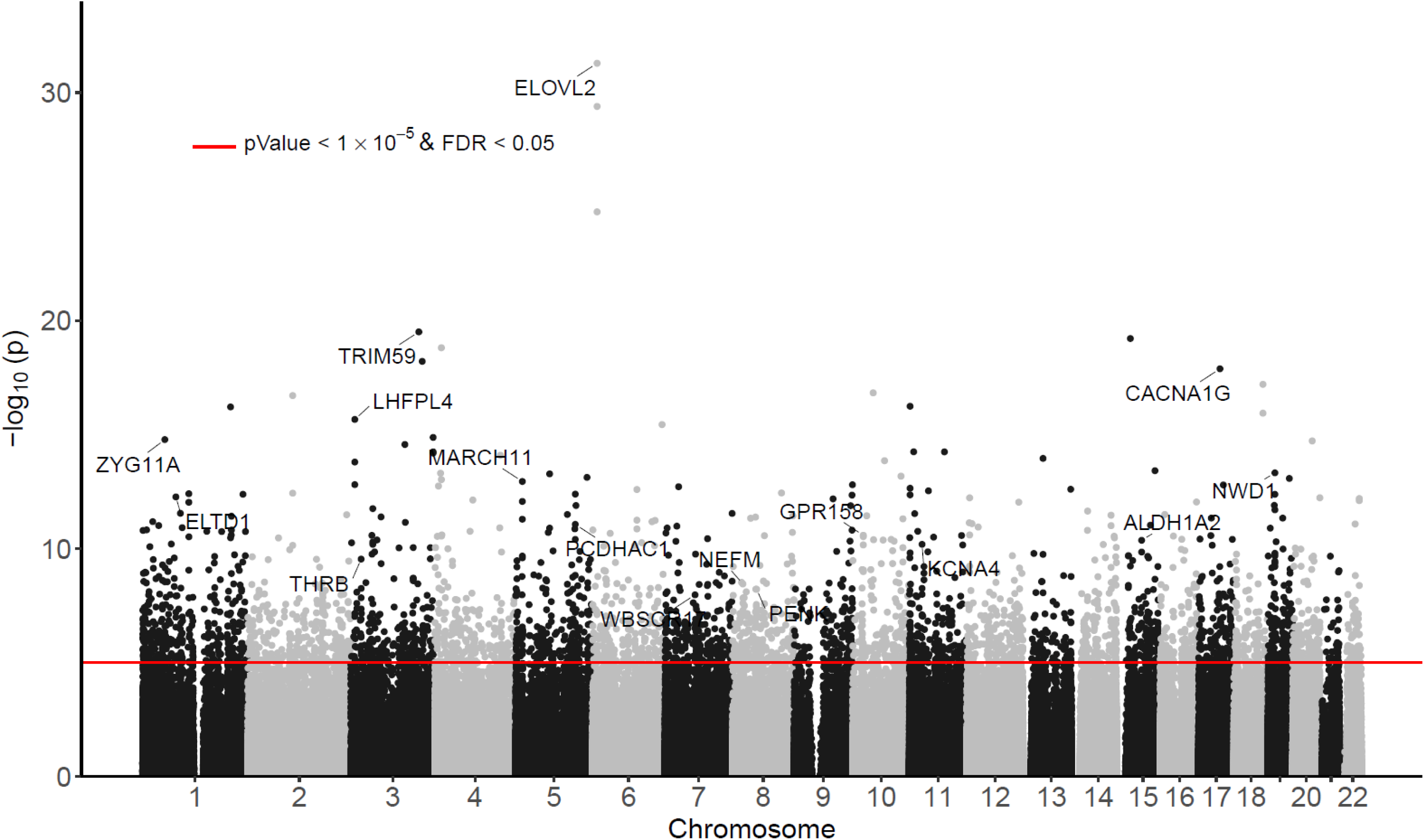
Manhattan plot of significant DNA methylation differences associated with chronological age in meta-analysis of FHS9 (FHS at exam 9) and ADNI datasets. The X-axis indicates chromosome number. The Y-axis shows –log_10_(*P-*value) of meta-analysis, with red line indicating significance threshold of 5% False Discovery Rate (FDR) and *P-*value < 10^-5^.

**Table 2.** Top 20 most significant CpGs associated with chronological age in meta-analysis of ADNI and FHS9 datasets. For each CpG, annotations include the location of the CpG (Chr, Position) and nearby genes based on GREAT software. The inverse-variance weighted meta-analysis regression model results include estimated effect size (estimate) where CpGs that are hyper-methylated with increased age have positive values, standard error (stdErr), *P-*value (pValue), and false discovery rate (fdr) accounting for multiple comparison corrections. The last column (direction) indicates the direction of effects in ADNI and FHS9 cohorts. All *P-*values are two-sided. In GREAT annotation, the numbers in parentheses indicate distance from the TSS. Highlighted in red text are promoter regions associated with the significant CpGs.

| CpG | Annotations |  |  | Meta-alysis results |  |  |  |  |
| --- | --- | --- | --- | --- | --- | --- | --- | --- |
|  | chr | position | GREAT_annotation | estimate | stdErr | pValue | fdr | direction |
| cg16867657 | chr6 | 11,044,643 | ELOVL2 (-331) | 0.033 | 0.003 | 5.17E-32 | 4.01E-26 | ++ |
| cg24724428 | chr6 | 11,044,654 | ELOVL2 (-342) | 0.026 | 0.002 | 4.03E-30 | 1.57E-24 | ++ |
| cg21572722 | chr6 | 11,044,660 | ELOVL2 (-348) | 0.016 | 0.002 | 1.71E-25 | 4.41E-20 | ++ |
| cg07553761 | chr3 | 160,450,188 | ENSG00000248710 (-361);TRIM59 (-361) | 0.027 | 0.003 | 3.14E-20 | 6.10E-15 | ++ |
| cg04875128 | chr15 | 31,483,691 | KLF13 (+156838);OTUD7A (+171647) | 0.028 | 0.003 | 6.13E-20 | 9.51E-15 | ++ |
| cg20816447 | chr4 | 15,479,156 | CC2D2A (+9293);FBXL5 (+176254) | -0.027 | 0.003 | 1.56E-19 | 2.02E-14 | -- |
| cg07323488 | chr3 | 168,467,524 | GOLIM4 (-371551);MECOM (+678780) | -0.021 | 0.002 | 6.16E-19 | 6.83E-14 | -- |
| cg11071401 | chr17 | 50,559,832 | CACNA1G (-1627) | 0.014 | 0.002 | 1.30E-18 | 1.27E-13 | ++ |
| cg13552692 | chr18 | 68,722,209 | CCDC102B (-75870);TMX3 (-6913) | -0.026 | 0.003 | 6.27E-18 | 5.41E-13 | -- |
| cg22796704 | chr10 | 48,465,490 | ARHGAP22 (+139463);MAPK8 (+158802) | -0.015 | 0.002 | 1.48E-17 | 1.15E-12 | -- |
| cg17268658 | chr2 | 105,399,287 | FHL2 (-65) | 0.017 | 0.002 | 1.95E-17 | 1.37E-12 | ++ |
| cg10332039 | chr11 | 1,827,434 | CTSD (-63444);SYT8 (-7009) | -0.020 | 0.002 | 5.75E-17 | 3.68E-12 | -- |
| cg16290275 | chr1 | 207,869,564 | CD34 (+41837);CD46 (+117509) | -0.019 | 0.002 | 6.16E-17 | 3.68E-12 | -- |
| cg19283806 | chr18 | 68,722,182 | CCDC102B (-75897);TMX3 (-6886) | -0.019 | 0.002 | 1.15E-16 | 6.40E-12 | -- |
| cg24866418 | chr3 | 9,552,397 | LHFPL4 (+1404);SETD5 (+154680) | 0.013 | 0.002 | 2.17E-16 | 1.12E-11 | ++ |
| cg20786223 | chr6 | 164,109,452 | QKI (+694811) | -0.021 | 0.003 | 3.64E-16 | 1.77E-11 | -- |
| cg11218872 | chr3 | 194,270,947 | CPN2 (+83320);HES1 (+134804) | -0.022 | 0.003 | 1.34E-15 | 6.10E-11 | -- |
| cg06784991 | chr1 | 52,843,095 | ZYG11A (+361) | 0.021 | 0.003 | 1.67E-15 | 7.19E-11 | ++ |
| cg07547549 | chr20 | 46,029,585 | SLC12A5 (+7870);NCOA5 (+60366) | 0.016 | 0.002 | 1.90E-15 | 7.75E-11 | ++ |
| cg12943155 | chr3 | 127,629,134 | PODXL2 (-46) | 0.025 | 0.003 | 2.73E-15 | 1.06E-10 | ++ |

In region-based analysis, after multiple comparisons correction, the comb-p software^14^ identified 629 significant DMRs associated with chronological age at a 5% Sidak-adjusted *P-*value. Among them, 556 DMRs were also identified by the coMethDMR software ^15^ at a 5% FDR (Supplementary Table 2, Table 3). These 556 DMRs contained an average of 4.3 CpGs per region. The majority of these DMRs (88.3%, 491 out of 556) were hypermethylated with increasing age, and most (90.3 %, 502 DMRs) were located within CpG islands or shores. Consistent with significant age-associated CpGs, among the 491 hypermethylated DMRs, the majority (75.2%, 369 DMRs) were found in gene promoter regions. On the other hand, among the 65 hypomethylated DMRs, most (56.9%, 37 DMRs) were located in distal regions.

**Table 3.**
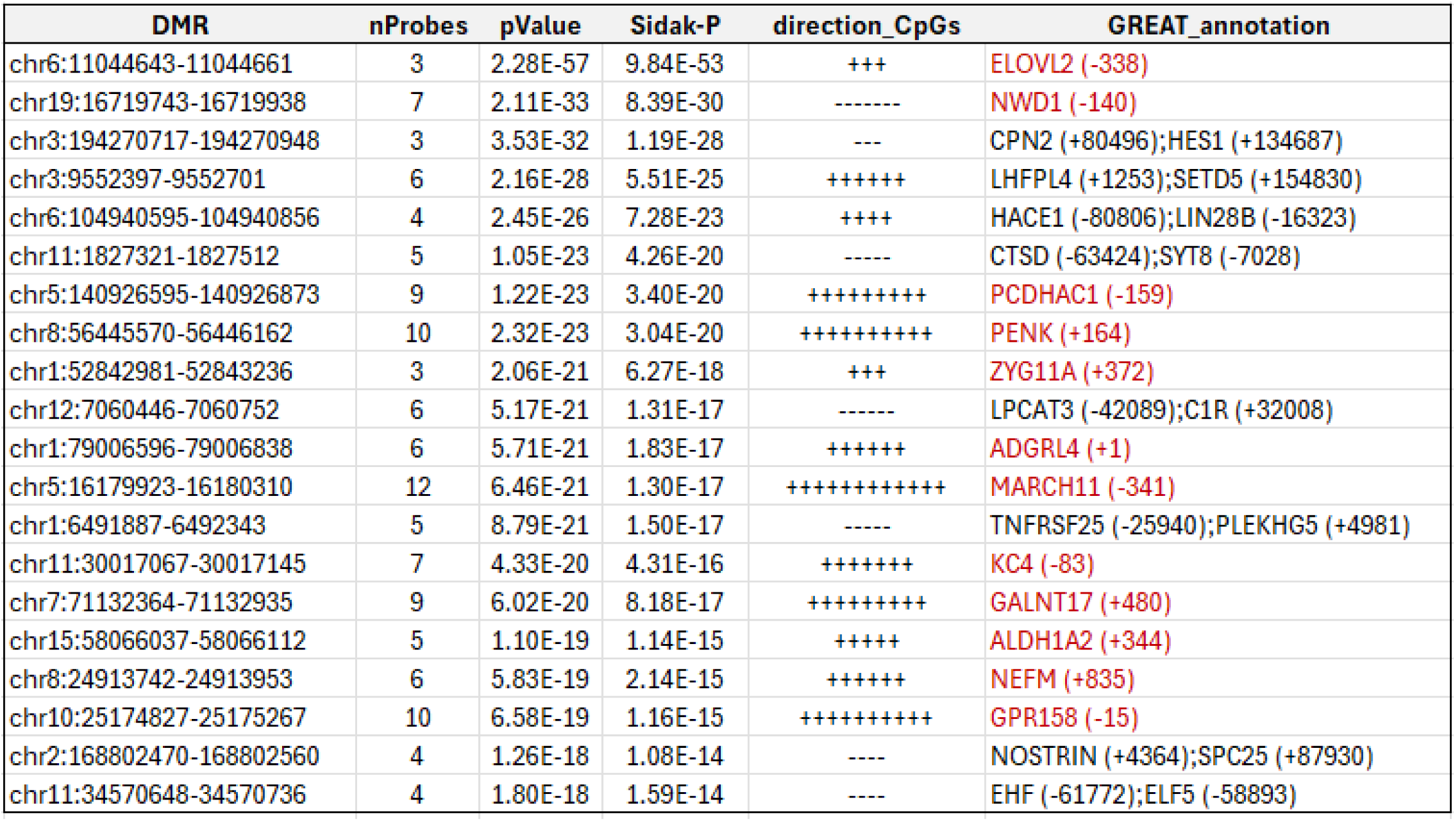
Top 20 most significant differentially methylated regions (DMRs) associated with chronological age identified by both comb-p and coMethDMR software in meta-analysis of ADNI and FHS9 datasets. For each DMR, annotations include location of the DMR (DMR) and nearby genes based on GREAT. Comb-p results include the number of probes (nProbes), nominal P-value (pValue), multiple comparison corrected P-value based on Sidak method (Sidak P), and direction of each CpG within the DMR (direction_CpGs). All P-values are two-sided. In GREAT annotation, the numbers in parentheses indicate distance from the TSS. Highlighted in red text are gene promoter regions associated with the DMRs.

### Correlation of significant DNAm with expression of nearby genes

To better understand the functional role of aging-associated DNAm, we overlapped the significant CpGs and DMRs with previously identified DNAm-to-RNA associations (eQTm), which were computed using paired blood DNA methylation and gene expression data collected from the same subjects in FHS.^16^ We identified 73 CpGs significantly correlated in *cis* (within 500 kb of the CpG) with target gene expression (Supplementary Table 3). More than half (62.1%, 64 out of 103) of these DNAm-to-RNA associations were negative. Among significant individual CpGs, the five most significant DNAm-to-RNA associations involved target genes *MX1, PDPR, MX2, SCP2*, and *ZNF418*. Similarly, among CpGs within DMRs, the five most significant DNAm-to-RNA associations involved target genes *SCP2, ZNF577*, and *ZNF418*.

*MX1* and *MX2* encode interferon-induced GTPases involved in immune response and antiviral defense pathways; dysregulation of these genes has been implicated in chronic inflammation associated with aging and AD ^17^. *PDPR* is involved in energy metabolism through regulation of pyruvate dehydrogenase activity, and is linked to mitochondrial dysfunction observed in aging and AD pathology ^18^. *SCP2* participates in lipid metabolism and cholesterol transport; these processes are critical in maintaining neuronal membrane integrity and potentially influencing amyloid-beta aggregation^19^. *ZNF577* and *ZNF418* are zinc finger transcription factors involved in gene regulation; altered expression of zinc finger proteins could disrupt transcriptional networks in aging brains and may contribute to AD pathology through epigenetic modulation^20^. These results highlight the functional relevance of aging-associated CpGs in regulating gene expression changes implicated in age-related inflammation, metabolism, and neurodegeneration.

### Pathway enrichment of aging-associated DNA methylation differences highlights biological hallmarks of aging and Alzheimer’s disease pathways

To better understand biological pathways enriched with significant aging-associated DNAm differences, we next performed pathway analysis using the methylGSA software^21^. At a 5% false discovery rate (FDR), we identified 26 KEGG pathways and 27 Reactome pathways significantly enriched with aging-associated DNAm (Supplementary Table 4, Table 4). Consistent with the 12 established biological hallmarks of aging^22^, these significant pathways highlight a number of critical molecular and cellular processes implicated in age-related functional decline. Several enriched pathways, such as *Type II diabetes mellitus*, *Integration of energy metabolism*, and *Peptide hormone metabolism*, are associated with the hallmark of deregulated nutrient sensing, which encompasses insulin/IGF-1 signaling and metabolic dysregulation in aging ^22^. Pathways associated with the altered intercellular communication hallmark include *Gap junction*, *Tight junction*, *Cell-adhesion molecules*, *ECM–receptor interaction*, and *Protein-protein interactions at synapses*, underscore that impaired cell-cell communication and structural remodeling accompany aging ^23^. Moreover, pathways such as the *Wnt signaling pathway* and *Hedgehog signaling pathway* are associated with the stem-cell exhaustion hallmark and altered developmental cues, highlighting aging-related deterioration in regenerative capacity and tissue homeostasis ^24^. Finally, mitochondrial dysfunction hallmark is represented by enrichment of the *Calcium signaling pathway* and *Purine metabolism*, which affect mitochondrial energy production and oxidative stress ^25^.

**Table 4.** The methylGSA software identified 26 KEGG pathways and 27 Reactome pathways significantly enriched with aging-associated CpGs at 5% FDR.

| KEGG pathways | Size | pValue | FDR |  | Reactome Pathway | Size | pValue | FDR |
| --- | --- | --- | --- | --- | --- | --- | --- | --- |
| Calcium signaling pathway | 167 | 1.00E-10 | 2.22E-08 |  | Neurotransmitter receptors and postsynaptic signal transmission | 194 | 8.20E-09 | 1.83E-05 |
| Dilated cardiomyopathy | 86 | 1.36E-07 | 1.51E-05 |  | Peptide ligand-binding receptors | 189 | 2.17E-08 | 2.42E-05 |
| Gap junction | 85 | 1.06E-05 | 7.87E-04 |  | Muscle contraction | 196 | 8.65E-08 | 4.83E-05 |
| Arrhythmogenic right ventricular card | 71 | 1.61E-05 | 8.91E-04 |  | Gastrulation | 121 | 8.65E-08 | 4.83E-05 |
| Axon guidance | 123 | 4.46E-05 | 1.98E-03 |  | Kidney development | 45 | 2.08E-07 | 9.27E-05 |
| Hypertrophic cardiomyopathy | 80 | 7.50E-05 | 2.54E-03 |  | GABA receptor activation | 57 | 8.64E-07 | 3.22E-04 |
| Cardiac muscle contraction | 68 | 1.02E-04 | 2.54E-03 |  | Neurexins and neuroligins | 50 | 2.01E-06 | 6.10E-04 |
| Protein digestion and absorption | 74 | 1.03E-04 | 2.54E-03 |  | Protein-protein interactions at synapses | 76 | 2.18E-06 | 6.10E-04 |
| Salivary secretion | 80 | 1.10E-04 | 2.54E-03 |  | Cardiac conduction | 127 | 4.94E-06 | 1.23E-03 |
| Cell adhesion molecules | 122 | 1.14E-04 | 2.54E-03 |  | Degradation of the extracellular matrix | 135 | 7.02E-06 | 1.57E-03 |
| Melanogenesis | 100 | 1.92E-04 | 3.84E-03 |  | G alpha (s) signalling events | 153 | 7.85E-06 | 1.59E-03 |
| Alzheimer disease | 143 | 2.07E-04 | 3.84E-03 |  | Opioid Signalling | 90 | 1.47E-05 | 2.73E-03 |
| Focal adhesion | 191 | 2.94E-04 | 5.02E-03 |  | Specification of the neural plate border | 17 | 2.36E-05 | 4.05E-03 |
| Type II diabetes mellitus | 47 | 6.00E-04 | 9.42E-03 |  | Amine ligand-binding receptors | 40 | 2.64E-05 | 4.21E-03 |
| Maturity onset diabetes of the young | 25 | 6.37E-04 | 9.42E-03 |  | Formation of the ureteric bud | 21 | 3.96E-05 | 5.89E-03 |
| Endocytosis | 196 | 1.63E-03 | 2.03E-02 |  | Potassium Channels | 102 | 4.74E-05 | 6.52E-03 |
| Bile secretion | 69 | 1.65E-03 | 2.03E-02 |  | Integration of energy metabolism | 105 | 4.97E-05 | 6.52E-03 |
| Regulation of actin cytoskeleton | 199 | 1.65E-03 | 2.03E-02 |  | Transport of inorganic cations/anions and amino acids/oligopeptides | 105 | 6.94E-05 | 8.61E-03 |
| Wnt signaling pathway | 146 | 2.50E-03 | 2.81E-02 |  | Cell junction organization | 105 | 1.35E-04 | 1.58E-02 |
| Tight junction | 125 | 2.53E-03 | 2.81E-02 |  | Cell-Cell communication | 140 | 1.42E-04 | 1.58E-02 |
| ECM-receptor interaction | 83 | 2.98E-03 | 3.15E-02 |  | Collagen formation | 88 | 1.73E-04 | 1.84E-02 |
| Amoebiasis | 102 | 3.13E-03 | 3.15E-02 |  | Regulation of beta-cell development | 40 | 1.85E-04 | 1.88E-02 |
| Basal cell carcinoma | 55 | 3.31E-03 | 3.20E-02 |  | Netrin-1 signaling | 42 | 2.01E-04 | 1.95E-02 |
| Long-term potentiation | 66 | 4.05E-03 | 3.75E-02 |  | Signaling by ERBB4 | 57 | 5.27E-04 | 4.90E-02 |
| Hedgehog signaling pathway | 55 | 4.72E-03 | 4.10E-02 |  | Peptide hormone metabolism | 83 | 5.63E-04 | 4.92E-02 |
| Purine metabolism | 150 | 4.80E-03 | 4.10E-02 |  | Molecules associated with elastic fibres | 37 | 5.72E-04 | 4.92E-02 |
|  |  |  |  |  | Incretin synthesis, secretion, and inactivation | 23 | 6.00E-04 | 4.96E-02 |

Importantly, several pathways enriched with aging-associated DNAm are directly relevant to AD. For example, the most significant KEGG pathway is the *Calcium signaling pathway*, which plays a central role in AD pathology by influencing amyloid-beta accumulation, tau phosphorylation, synaptic dysfunction, and neuronal death ^26^. Pathways related to neuronal communication and synaptic function, including *Neurotransmitter receptors and postsynaptic signal transmission*, *Long-term potentiation*, and *Protein-protein interactions at synapses*, are also closely linked to cognitive impairment observed in AD ^27^. Additionally, the *Wnt signaling pathway* has been implicated in AD through its influence on synaptic plasticity, neuronal survival, and amyloid-beta production ^28^. *ECM–receptor interaction*, *Cell adhesion molecules*, and *Focal adhesion* pathways have roles in neuronal connectivity and integrity, processes often compromised during AD pathogenesis ^29^. Moreover, enrichment of the *Type II diabetes mellitus* and *Integration of energy metabolism* pathways supports the hypothesis of AD as a metabolic disease ^30^, consistent with the increased recognition that metabolic dysfunction contributes to AD pathology. Notably, the KEGG pathway *Alzheimer’s disease* is significantly enriched with aging-associated CpGs (*P-*value = 2.07×10^-4^, FDR = 0.0038). Collectively, these pathways enriched with aging-associated DNAm highlight multiple biological processes linked to AD pathology, suggesting that age-related epigenetic changes may contribute significantly to the disease.

### Co-localization analysis nominated genetic variants associated with both dementia and aging-associated DNA methylation

To further understand the functional roles of the aging-associated CpGs and DMRs in AD, we performed integrative analyses of aging-associated DNAm with genetic data. First, to identify methylation quantitative trait loci (mQTLs) for the significant aging-associated DNAm differences, we searched the GoDMC database^31^. Among the 3758 aging-associated CpGs (Supplementary Table 1) and the 2409 CpGs located in significant DMRs (Supplementary Table 2), we found that 1696 CpGs (45.1%) had 311,054 mQTLs (Supplementary Table 5). This proportion is consistent with findings from a recent large mQTL meta-analysis in blood, which estimated that approximately 45% of CpGs on the Illumina array are influenced by genetic variants^31^.

As the aging processes and dementia may share common genetic factors, we next evaluated whether these mQTLs overlapped with genetic risk loci implicated in dementia, by comparing them with the genetic variants identified in a recent ADRD (Alzheimer’s and related dementia) meta-analysis. ^32^ While no mQTLs overlapped with genome-wide significant loci (*P-*value< 5 ×10^-8^) for ADRD, we found 1168 mQTLs overlapped with genetic variants reaching a suggestive significance threshold at *P* < 1×10^-5^ (Supplementary Table 6).

Given the observed overlap between mQTLs and ADRD genetic risk loci, we sought to determine whether the association signals at these loci (variant to CpG methylation levels and variant to ADRD status) were driven by a single shared causal variant or by distinct variants in proximity. To this end, we performed a co-localization analysis using the method described in Giambartolomei et al. (2014)^33^. The results provided strong evidence ^34^ (PP3+PP4 > 0.90, PP4 > 0.8 and PP4/PP3 > 5) supporting a shared causal variant in 32 genomic regions influencing both traits. Among the associated CpGs, several were located in promoter regions of genes including *ABI3, RELN, ZNF233, DLG4, HIVEP3, DNTT, HSD3B7, RNF39, PPM1E,* and *HSD3B7* (Supplementary Table 7).

### Aging-associated DNAm differences are significantly associated with Alzheimer’s disease in independent studies

To further investigate the relevance of aging-associated DNAm differences to AD, we next examined their presence in previous AD-related DNAm studies using our recently developed MIAMI-AD database (https://miami-ad.org/)^35^. We considered AD phenotypes including clinical diagnoses of mild cognitive impairment (MCI), AD, or dementia; AD-related neuropathology in brain tissue; and cerebrospinal fluid (CSF) AD biomarkers. To ensure independence from our aging study, we excluded datasets that used samples from the ADNI or FHS studies.

Applying a stringent Bonferroni correction for 5362 CpGs (including 3758 significant individual CpGs and 1604 CpGs located within DMRs, at a significance threshold of *P-*value < 9.32 ×10^-6^), we identified 33 CpGs that were also significant in independent studies of AD. These studies analyzed brain tissue from the prefrontal cortex (PFC), temporal cortex (TC), parahippocampal gyrus (PHG), or middle temporal gyrus (MTG) (Supplementary Table 8). Notably, all these external studies adjusted for age and other covariates, so the observed DNAm-to-AD associations are independent of age effects.

These 33 CpGs were located in the promoter regions of *ACADS, CHST9, DIO3, EDARADD, GP5, GPR56, HTR4, LIMD1*, *PENK, SLC24A3* genes, and other genomic regions. At the more relaxed nominal significance threshold (*P-*value < 0.05), we found that 1846 (34.4%) of the 5362 aging-associated CpGs were associated with AD phenotypes in prior studies. Among them, 1159 (62.8%) were identified in brain-based DNAm studies, 503 (27.2%) in blood-based studies, and 184 (10.0%) were observed in both blood and brain DNAm studies of AD. The higher number of overlapping CpGs in brain studies may reflect both the greater number of brain-based AD studies compared to blood-based studies included in the MIAMI-AD database, and the larger biological variability in blood-based DNAm data ^36^, which can reduce statistical power relative to brain-based studies.

Limiting to blood-based studies, we next compared the direction of methylation changes in aging and AD among the 687 CpGs (503 + 184 CpGs), we found less than half (278 CpGs, 40.5%) showed concordant changes: 81 CpGs were hypomethylated and 197 CpGs were hypermethylated in both aging and AD. The remaining 409 CpGs (59.5%) were discordant: 45.4% of the CpGs (312 CpGs) were hypermethylated in aging but hypomethylated in AD, and the remaining 14.1% CpGs (97 CpGs) showed hypomethylation in aging but hypermethylation in AD.

At a 5% FDR, enrichment analysis showed that CpGs with concordant DNAm changes in aging and AD were significantly over-represented in the *phasic smooth muscle contraction* pathway, reflecting vascular dysfunction common in AD, as well as in the developmental pathways *tripartite regional subdivision* and *anterior/posterior axis specification in embryo*. On the other hand, CpGs showing discordant DNAm changes between aging and AD were significantly enriched in *neuroactive ligand signaling* and *neuron migration* pathways, key regulators of neuronal communication and synaptic plasticity that are dysregulated in AD (Supplementary Table 9).

### Brain-to-blood DNAm correlation analysis identified aging-associated blood DNAm with concordant cross-tissue changes

To identify aging-associated DNA methylation (DNAm) with the potential to serve as biomarkers, we evaluated brain-to-blood DNA methylation correlations of the aging-associated DNAm. To this end, we utilized the London cohort dataset, which included 69 pairs of matched brain and blood samples^37^, and computed the Spearman correlations between brain and blood DNA methylation levels, after removing the effects of estimated cell type proportions, batch effects, age, and sex in brain and blood samples separately (Methods).

Among the 3758 significant individual CpGs associated with aging and 1604 CpGs located in aging DMRs, DNAm at 23 CpGs showed significant brain-to-blood correlations (*FDR* < 0.05) (Supplementary Table 10). All 23 CpGs showed a significant positive association, ranging from 0.423 to 0.626. These CpGs were located in the promoter of *CNTNAP2, PODXL2, MARCH11, OCIAD2, SCGN, ZNF233, C3orf18, ELOVL2, ZNF442, PDE1B* genes and other genomic regions. The consistent cross-tissue methylation patterns at these CpGs may indicate important shared regulatory roles during aging.

By intersecting these 23 CpGs with the 1846 CpGs significantly associated with both aging and AD described above, we identified 9 CpGs, located in the promoter regions of *PDE1B*, *ELOVL2*, *PODXL2* genes and other genomic regions, that showed both strong concordance in brain-to-blood DNAm levels, as well as association with AD diagnosis or AD neuropathology in independent studies (Supplementary Table 11). One notable example is cg26019680 in the promoter of *PODXL2*, which showed high correlation of DNAm levels between blood and four brain regions (prefrontal cortex, entorhinal cortex, superior temporal gyrus, and cerebellum) (Supplementary Figure 1). This CpG is part of a hypermethylated DMR associated with aging and also displays hypermethylation in male AD cases ^38^. Such CpGs represent promising candidates for future biomarker development.

## DISCUSSION

We performed a comprehensive meta-analysis of two large, independent, blood-based DNAm datasets generated by the FHS and ADNI studies, which were measured using the same Infinium MethylationEPIC BeadChip. To characterize DNAm changes associated with chronological aging, we analyzed samples collected from participants older than 65 years at the time of blood collection who remained dementia-free throughout follow-up in both cohorts. To minimize false positives, we applied two complementary methods to identify DMRs, selecting only those genomic regions significantly associated with chronological age in both analyses after multiple-testing correction, and have consistent directionality across all CpGs within each region. To explore molecular intersections between normal aging and dementia, we integrated our findings with multi-omics data from recent large-scale studies, including expression quantitative trait methylation, methylation quantitative trait loci, ADRD GWAS summary statistics, brain-to-blood DNAm correlations, and AD-associated DNAm studies manually curated in the MIAMI-AD database.

At a 5% FDR, we identified 3758 CpGs and 556 DMRs consistently associated with chronological age in both the FHS and ADNI datasets. Our findings are consistent with previous studies of the aging epigenome. ^39^ Notably, most of the hypermethylated CpGs were located within promoter regions, which have previously been reported to undergo epigenetic silencing with age ^3,40,41^. In contrast, hypomethylated CpGs predominantly mapped to distal genomic regions, consistent with prior observations of global hypomethylation in intergenic areas during aging ^42^. These patterns were further supported by our region-based analysis, in which 88.3% of DMRs showed age-associated hypermethylation, with the vast majority (91.0%) located in gene promoter regions. However, it is important to note that the results of this study are limited to the probe content of the Illumina EPIC array. Future studies using sequencing-based technology may offer a more comprehensive view of the aging methylome.

Importantly, a review of recent literature revealed that many of the aging-associated CpGs and DMRs we identified in this study have previously been implicated in AD. Among the top 20 CpGs (Table 2), the three most significant CpGs were located in the *ELOVL2* gene, whose hypermethylation has consistently been demonstrated to be a robust epigenetic biomarker of chronological aging across multiple tissues, including brain ^43^, blood ^44^, and saliva ^45^. Additionally, the EpiAge clock, built solely on these three CpGs (cg16867657, cg21572722, and cg24724428) shows significant acceleration in individuals with mild cognitive impairment (MCI) compared to controls. ^45^ A recent GWAS also identified a genetic variant in *ELOVL2* significantly associated with an increased risk of AD, potentially through alterations in lipid metabolism. ^46^ *TRIM59* is another gene that shows age-associated hypermethylation. A recent study reported hypermethylation of *TRIM59* in blood samples from AD patients compared to healthy controls. ^47^ This hypermethylation was correlated with abnormalities in DNA repair and cell cycle regulation, two critical processes involved in AD pathology. *CACNA1G* encodes a calcium channel, and its expression decreases with age in both human and mouse brains, a decline further exacerbated in AD. This downregulation may disrupt calcium homeostasis, promote amyloid-beta production, and contribute to cognitive decline. ^48^ Finally, *FHL2* gene is also hypermethylated with age, it regulates inflammatory responses and adipose tissue metabolism, both of which are increasingly recognized as important contributors to AD pathophysiology.^49^

Similarly, among the top 20 most significant DMRs (Table 3), *PCDHAC1* is a member of the protocadherin family involved in synapse formation and stabilization, processes whose disruption has also been implicated in AD pathology. ^50^ The *GALNT17* gene encodes an enzyme in the GALNT family, which initiates glycosylation (the addition of sugar molecules to proteins), a process recently linked to microglial-driven neuroinflammation and exacerbation of AD pathology in the brain. ^51^ Also, the *ALDH1A2* gene encodes a key enzyme in retinoic acid synthesis, a pathway known to support neuroplasticity and memory. Reduced *ALDH1A2* expression has been observed in multiple AD mouse models, even at early disease stages. This decline may play an initiating role in AD pathogenesis, and pharmacologic restoration of RA signaling has shown therapeutic promise ^52^. Another noteworthy gene is *NEFM* (Neurofilament Medium Polypeptide). In AD mouse models, elevated *NEFM* expressions were associated with neuronal dysfunction, including axonal damage and disrupted cytoskeletal integrity. Notably, reducing *NEFM* gene expression through IGF1R inhibition provided neuroprotection, leading to improved neuronal function and reduced neuroinflammation. ^53^

Finally, *GPR158* encodes a G protein-coupled receptor predominantly localized to neurons in the cortical and hippocampal regions, areas critical for synaptic architecture and plasticity, which are disrupted in AD. In a recent functional genomics study ^54^, *GPR158* was identified as one of five hub genes significantly downregulated in the temporal cortex of AD patients. Gene ontology analysis revealed that these hub genes, including *GPR158*, are enriched in pathways related to synaptic function and memory processes, suggesting that their reduced expression may contribute to synaptic failure in AD. Notably, *GPR158* expression was inversely correlated with β-secretase (BACE1) activity in AD brain samples, indicating that lower *GPR158* levels are associated with increased BACE1 activity, which could in turn enhance Aβ production from Amyloid Precursor Protein (APP).

In addition to corroborating previous findings, our analyses also identified several novel differentially methylated genes that have potential implications in AD. For instance, in the comparison of aging-associated CpGs with those reported in prior AD studies, cg02336827, located in the promoter region of the *LIMD1* gene and hypomethylated with age, emerged as one of the most significant CpGs, showing a *P-* value of 4.63×10^-7^ and consistent hypomethylation in AD (Supplementary Table 8).^55^ *LIMD1* encodes a scaffold protein involved in key cellular functions, including transcriptional repression, microRNA-mediated gene silencing, and cytoskeletal organization. Although *LIMD1* itself has not been directly implicated in AD, other LIM domain-containing proteins have been linked to neurodegenerative processes. Notably, LIM kinase 1 (*LIMK1*), which regulates actin cytoskeleton dynamics, has been associated with synaptic dysfunction in AD. ^56^ These findings suggest that *LIMD1* may represent a novel candidate for future AD studies.

To reveal additional biological insights linking age-associated blood methylation to dementia risk, we performed several additional integrative analyses. Our pathway enrichment analysis revealed that aging-associated CpGs were significantly overrepresented in KEGG and Reactome pathways related to *calcium signaling*, *Wnt/Hedgehog signaling*, *extracellular matrix (ECM)* and *cell adhesion*, and *metabolic regulation*, systems repeatedly implicated in AD pathophysiology (Table 4). Colocalization analyses identified 32 loci where the same causal variant likely influences both DNAm and ADRD risk. Notably, shared signals which harbored both mQTLs and AD GWAS hits were observed at *ABI3*, *RELN*, and *DLG4,* genes involved in microglial activation, synaptic organization, and neuronal plasticity. ^57-59^

Furthermore, we found that approximately one-third of aging-associated CpGs were also associated with AD or AD neuropathology in independent AD DNAm studies after adjusting for age and other covariates, with about 40% displaying concordant direction of effect (Supplementary Table 8). Divergent patterns (e.g., hypermethylation with aging but hypomethylation in AD) may reflect a dysregulation of epigenetic programs in normal aging, whereby protective or adaptive methylation changes that typically accumulate with age are either not established or are actively reversed in AD. This supports the model proposed by Berger and colleagues that AD is not merely an acceleration of aging but also involves a disruption of homeostatic epigenetic regulation that contributes to neurodegeneration ^60^. Taken together, our results suggest that a number of age-related epigenetic changes in peripheral blood function not only as biomarkers of aging, but also reflect relevant changes in AD or AD neuropathology, and maybe influenced by genetic risk variants for AD.

We further demonstrated that 23 aging-associated CpGs showed significant positive correlations between blood and brain frontal cortex (Supplementary Table 10). Among the genes associated with these CpGs, the *CNTNAP2* gene encodes a cell adhesion molecule involved in various critical neuronal functions, including axonal organization, synaptic regulation, and neuronal migration. Recent studies have identified genetic variants in the *CNTNAP2* gene significantly associated with AD ^61^, and altered *CNTNAP2* expression levels have been observed in the brains of AD patients ^62^. Also, the *SCGN* gene encodes the calcium-binding protein secretagogin. Importantly, *SCGN* has been postulated to have neuroprotective effects against neurodegeneration, as neurons expressing *SCGN* were largely resistant to cell death in human hippocampus. ^63^ Moreover, 9 of the 23 CpGs, including loci in *ELOVL2, PODXL2,* and *PDE1B*, were also significantly associated with AD or AD neuropathology in independent datasets, after adjusting for age and other covariates (Supplementary Table 11). *ELOVL2*, in particular, is a well-established component of several epigenetic clocks ^3,41,64^ and has been linked to both neuronal lipid homeostasis and cognitive decline ^65^. Prospective evaluation of these CpGs in longitudinal cohorts will clarify their utility for early dementia risk stratification.

The strengths of this study include the careful selection of samples, rigorous harmonization and quality control procedures, and robust analytical approaches applied to two well-characterized cohorts. In addition to meta-analyses of individual CpGs and genomic regions, we also performed comprehensive integrative analyses that incorporated information from eQTMs, mQTLs, ADRD GWAS, brain-to-blood DNAm correlations, pathway enrichment, and the manually curated MIAMI-AD database.

Several limitations of this study are in order. First, the DNAm was measured in whole blood, which may not fully capture cell-type-specific changes. Future work leveraging single-cell technology will offer more insight into the specific cell types affected by the aging-associated DNAm differences discovered in this study. Second, given that brain and blood derive from distinct cell lineages, blood-based DNAm markers may not accurately reflect brain-specific methylation changes. To address this, we prioritized DNAm changes with concordant brain-to-blood associations using a large publicly available DNAm dataset with matched brain and blood DNA methylation levels. Third, both the ADNI and FHS cohorts are predominantly of European ancestry, limiting generalizability to more diverse populations. Fourth, we analyzed FHS samples collected at Exam 9, and the earliest ADNI visit with available DNAm data; thus, the cross-sectional nature of our analysis precludes causal inference. Large independent longitudinal cohorts with incident dementia outcomes are needed to validate and prioritize the DNAm differences identified here as potential early markers of disease onset.

In summary, our results reveal that hypermethylation within gene promoters and hypomethylation in distal regions form a dominant aging signature in peripheral blood. These aging-associated DNAm differences converges on immune, metabolic, and synaptic pathways implicated in AD. We further prioritized a number of CpGs where DNAm is influenced by mQTLs colocalizing with dementia risk loci, or show strong blood-to-brain concordance and are associated with AD in independent studies adjusting for age and other covariates, highlighting their promise as blood-based AD biomarkers in future research.

## METHODS

### Study datasets

Our meta-analysis included 475 blood DNA methylation (DNAm) samples generated by two independent studies: the ADNI and the FHS Offspring cohort study. In ADNI, we analyzed each participant’s earliest visit with available DNAm data, and for the FHS Offspring cohort, we used DNAm data from Exam 9.

### Pre-processing of DNA methylation data

Supplementary Table 12 shows the number of CpGs and samples at each quality control (QC) step. The pre-processing procedures were previously described elsewhere ^8^. Briefly, for each dataset, the QC of probes involved selecting probes with a detection *P-*value < 0.01 in 90% or more of the samples, probes that start with “cg”, and removing probes that are located on X and Y chromosomes, are cross-reactive ^66^, or located close to single nucleotide polymorphism (SNPs). The QC of samples included removing samples with bisulfite conversion rate lower than 85%, as well as samples for which the DNAm predicted sex status differed from the recorded sex status. In addition, we performed principal component analysis (PCA) using the 50,000 most variable CpGs to identify outliers. Samples outside the range of ±3 standard deviations from the mean of PC1 and PC2 were excluded. The quality-controlled data was next normalized using the dasen method, as implemented in the wateRmelon R package ^67^. To correct batch effects from methylation plates, we used the BEclear R package ^68^.

Immune cell type proportions (B lymphocytes, natural killer cells, CD4+ T cells, CD8+ T cells, monocytes, neutrophils, and eosinophils) were estimated using the EpiDISH R package ^69^. Granulocyte proportions were computed as the sum of neutrophil and eosinophil proportions^36,38,70^, since both neutrophils and eosinophils are classified as granular leukocytes.

For both studies, we included samples from self-reported non-Hispanic white individuals over 65 years of age. To avoid inflation due to family structure in FHS9, we chose only one individual per family, prioritizing the sample with the highest bisulfite conversion rate. In ADNI and FHS9, the participants were followed for up to 11.11 and 7.72 years after their blood draw, respectively. We excluded samples from individuals who developed dementia at the time of blood draw or during the follow-up period.

### Association of DNA methylation at individual CpGs with dementia

For each dataset, the association between CpG methylation levels and chronological age at blood draw was assessed using linear statistical models. Given that methylation *M-*values (logit transformation of methylation beta values) have better statistical properties (i.e., homoscedasticity) for linear regression models ^71^, we used the *M-*values as the outcome variable in our statistical models. For both the ADNI and FHS9 datasets, we adjusted for potential confounding factors including sex and estimated major immune cell-type proportions in the samples. The linear model we used is methylation M value ∼ age + sex + immune cell-type proportions (B, NK, CD4T, Mono, Gran).

### Inflation assessment and correction

Genomic inflation factors (lambda values) were estimated using both the conventional approach ^72^ and the *bacon* method ^73^, which was proposed specifically for EWAS. Using the conventional approach, the estimated λ values were 1.097 for ADNI and 1.229 for FHS9. The inflation factors estimated by the bacon approach (λ.bacon) were 1.033 and 1.136 for the ADNI and FHS9 datasets, respectively. The estimated bias from the bacon method were -0.048 for ADNI and -0.044 for FHS9.

After genomic correction using the bacon method^73^, as implemented in the bacon R package, the estimated bias were -1.35×10^-4^ and 4.70×10^-4^, and the estimated inflation factors were λ = 1.049 and 1.063, and λ.bacon = 1.002 and 1.006 for the ADNI and FHS9 and datasets, respectively. The bacon method was then used to compute bacon-corrected effect sizes, standard errors, and *P-*values for each dataset.

### Meta-analysis

To meta-analyze individual CpG results across both the FHS9 and ADNI datasets, we used the inverse-variance weighted fixed-effects model^74^, as implemented in the meta R package. To correct for multiple comparisons, we computed the false discovery rate (FDR). We considered CpGs with an FDR less than 5% in meta-analysis of the FHS9 and ADNI datasets, with a consistent direction of change in estimated effect sizes, and a nominal *P-*value less than 1×10^-5^ as statistically significant.

### Differentially methylated regions analysis

To identify differentially methylated regions (DMRs) significantly associated with chronological age, we used two analytical approaches, the comb-p ^14^ approach and the coMethDMR ^15^ approach, and selected significant DMRs identified by both methods. Briefly, comb-p takes single CpG *P-*values and locations of CpG sites to scan the genome for regions enriched with a series of adjacent low *P-*values. In our analysis, we used meta-analysis *P*-values of the two blood sample cohorts as input for comb-p. We used the default parameter setting for our comb-p analysis, with parameters --seed 1e-3 and --dist 200, which required a *P-* value of 10^-3^ to start a region and extend the region if another *P-*value was within 200 base pairs. As comb-p uses the Sidak method ^75^ to adjust for multiple comparisons, we selected DMRs with Sidak *P-*values less than 0.05. In addition, we further required the selected DMRs to have at least 3 CpGs and a consistent direction of change across all CpGs mapped within the region.

In the coMethDMR approach, the “contiguous genomic regions” are genomic regions on the Illumina array covered with clusters of contiguous CpGs where the maximum separation between any two consecutive probes is 200 base pairs. First, coMethDMR selects co-methylated sub-regions within the contiguous genomic regions. Next, we summarized methylation M values within these co-methylated sub-regions using medians and tested them against chronological age at blood draw, adjusting for sex, and estimated immune cell-type proportions using linear regression models. The bacon method ^73^ was next applied to cohort-specific coMethDMR test statistics to obtain inflation-corrected effect sizes, standard errors, and *P-*values, which were then combined by inverse-variance weighted meta-analysis models using R package meta. We considered co-methylated DMRs with at least 3 CpGs, with a consistent direction of change in estimated effect sizes in both datasets, and a meta-analysis FDR < 0.05 to be significant in the coMethDMR analysis. Finally, we selected significant DMRs identified by both comb-p and coMethDMR approaches for subsequent analyses.

### Functional annotation of significant methylation associations

Significant methylation at individual CpGs and DMRs was annotated using both the Illumina (UCSC) gene annotation and Genomic Regions Enrichment of Annotations Tool (GREAT) software ^76^, which associates genomic regions with target genes.

### Pathway analysis

To identify biological pathways enriched with significant DNA methylation differences, we used the methylRRA function in the methylGSA R package ^21^, which used *P*-values from the meta-analysis of FHS9 and ADNI datasets as input. Briefly, methylGSA first computes a gene-wise *ρ* value by aggregating *P*-values from multiple CpGs mapped to each gene. Next, the different number of CpGs on each gene is adjusted by Bonferroni correction. Finally, a Gene Set Enrichment Analysis ^77^ (in pre-rank analysis mode) is performed to identify pathways enriched with significant CpGs. We analyzed pathways in the KEGG and REACTOME databases. Pathways with FDR less than 0.05 were considered to be statistically significant.

### Integrative analyses with gene expression, genetic variants, and brain-to-blood correlations

To evaluate the effect of DNA methylation on the expression of nearby genes, we overlapped our aging-associated CpGs, including both significant individual CpGs and those located within DMRs, with eQTm analysis results in Supplementary Tables 2 and 3 of Yao et al. (2021)^16^.

For correlation and overlap of aging-associated CpGs with genetic susceptibility loci, we searched the GoDMC database.^17^ To select significant blood mQTLs of aging-associated CpGs in GoDMC, we used the same criteria as the original study, ^31^ that is, considering a cis *P-*value smaller than 10^-8^ and a trans *P-*value smaller than 10^-14^ as significant. The genome-wide summary statistics for genetic variants associated with dementia described in Bellenguez et al. (2022) ^32^ were obtained from the European Bioinformatics Institute GWAS Catalog under accession no. GCST90027158. Colocalization analysis was performed using the coloc R package.

To assess the correlation of aging-associated CpGs and DMRs methylation levels in blood and brain samples, we used the London dataset, which consisted of 69 samples with matched PFC and blood samples^37^. We assessed the association of brain and blood methylation levels at aging-associated CpGs by performing an adjusted correlation analysis using methylation residuals (*r_resid_*), in which we first removed the effects of estimated neuron proportions in brain samples (or estimated immune cell-type proportions in blood samples), array, age at death for brain samples (or age at blood draw for blood samples), and sex from DNA methylation *M*-values.

### Validation using independent datasets

To compare our results with previous independent AD studies, we searched aging-associated CpGs (both significant individual CpGs and those located in DMRs) using the CpG Query tool in the MIAMI-AD database^78^. For input on phenotype, we selected “AD Biomarker”, “AD Neuropathology”, “Dementia Clinical Diagnosis”, and “Mild Cognitive Impairment”. Only studies external to the ADNI and FHS datasets were included.

## Data availability

The ADNI and Framingham Heart Study datasets can be accessed from http://adni.loni.usc.edu and the dbGap database (accession: phs000974.v5.p4).

## Code availability

The scripts for the analyses performed in this study are at https://github.com/TransBioInfoLab/AD-Aging-blood-sample-analysis.

## Data Availability

https://www.ncbi.nlm.nih.gov/projects/gap/cgi-bin/study.cgi?study_id=phs000974.v6.p5

## Acknowledgment

This research was supported by US National Institutes of Health grants R61NS135587 (L.W.), RF1NS128145 (L.W.), and R01AG062634 (E.R.M, B.W.K., L.W.).

Data used in preparation of this article were obtained from the Alzheimer’s Disease Neuroimaging Initiative (ADNI) database (adni.loni.usc.edu). As such, the investigators within the ADNI contributed to the design and implementation of ADNI and/or provided data but did not participate in analysis or writing of this report. A complete listing of ADNI investigators can be found at: http://adni.loni.usc.edu/wp-content/uploads/how_to_apply/ADNI_Acknowledgement_List.pdf Data collection and sharing for the ADNI dataset was funded by the Alzheimer’s Disease Neuroimaging Initiative (ADNI) (National Institutes of Health Grant U01 AG024904) and DOD ADNI (Department of Defense award number W81XWH-12-2-0012). ADNI is funded by the National Institute on Aging, the National Institute of Biomedical Imaging and Bioengineering, and through generous contributions from the following: AbbVie, Alzheimer’s Association; Alzheimer’s Drug Discovery Foundation; Araclon Biotech; BioClinica, Inc.; Biogen; Bristol-Myers Squibb Company; CereSpir, Inc.; Cogstate; Eisai Inc.; Elan Pharmaceuticals, Inc.; Eli Lilly and Company; EuroImmun; F. Hoffmann-La Roche Ltd and its affiliated company Genentech, Inc.; Fujirebio; GE Healthcare; IXICO Ltd.; Janssen Alzheimer Immunotherapy Research & Development, LLC.; Johnson & Johnson Pharmaceutical Research & Development LLC.; Lumosity; Lundbeck; Merck & Co., Inc.; Meso Scale Diagnostics, LLC.; NeuroRx Research; Neurotrack Technologies; Novartis Pharmaceuticals Corporation; Pfizer Inc.; Piramal Imaging; Servier; Takeda Pharmaceutical Company; and Transition Therapeutics. The Canadian Institutes of Health Research is providing funds to support ADNI clinical sites in Canada. Private sector contributions are facilitated by the Foundation for the National Institutes of Health (www.fnih.org). The grantee organization is the Northern California Institute for Research and Education, and the study is coordinated by the Alzheimer’s Therapeutic Research Institute at the University of Southern California. ADNI data are disseminated by the Laboratory for Neuro Imaging at the University of Southern California.

## Author Contributions

L.W., J.Y., E.R.M., and W.Z. designed the computational analyses. W.Z., D.L., L.G., M.A.S., and L.W. analyzed the data. L.W., J.Y., E.R.M, X.C., and B.W.K. contributed to the interpretation of the results. L.W., W.Z. wrote the paper, and all authors participated in the review and revision of the manuscript. L.W. conceived the original idea and supervised the project.

## Competing Interest

The authors declare that they have no conflict of interest.

## Notes

### Competing Interest Statement

The authors have declared no competing interest.

### Author Declarations

IRB of University of Miamii gave ethical approval for this work

