## Supplementary material for "The Aging Epigenome: Integrative Analyses Reveal Functional Overlap with Alzheimer’s Disease": Supp Figure 1

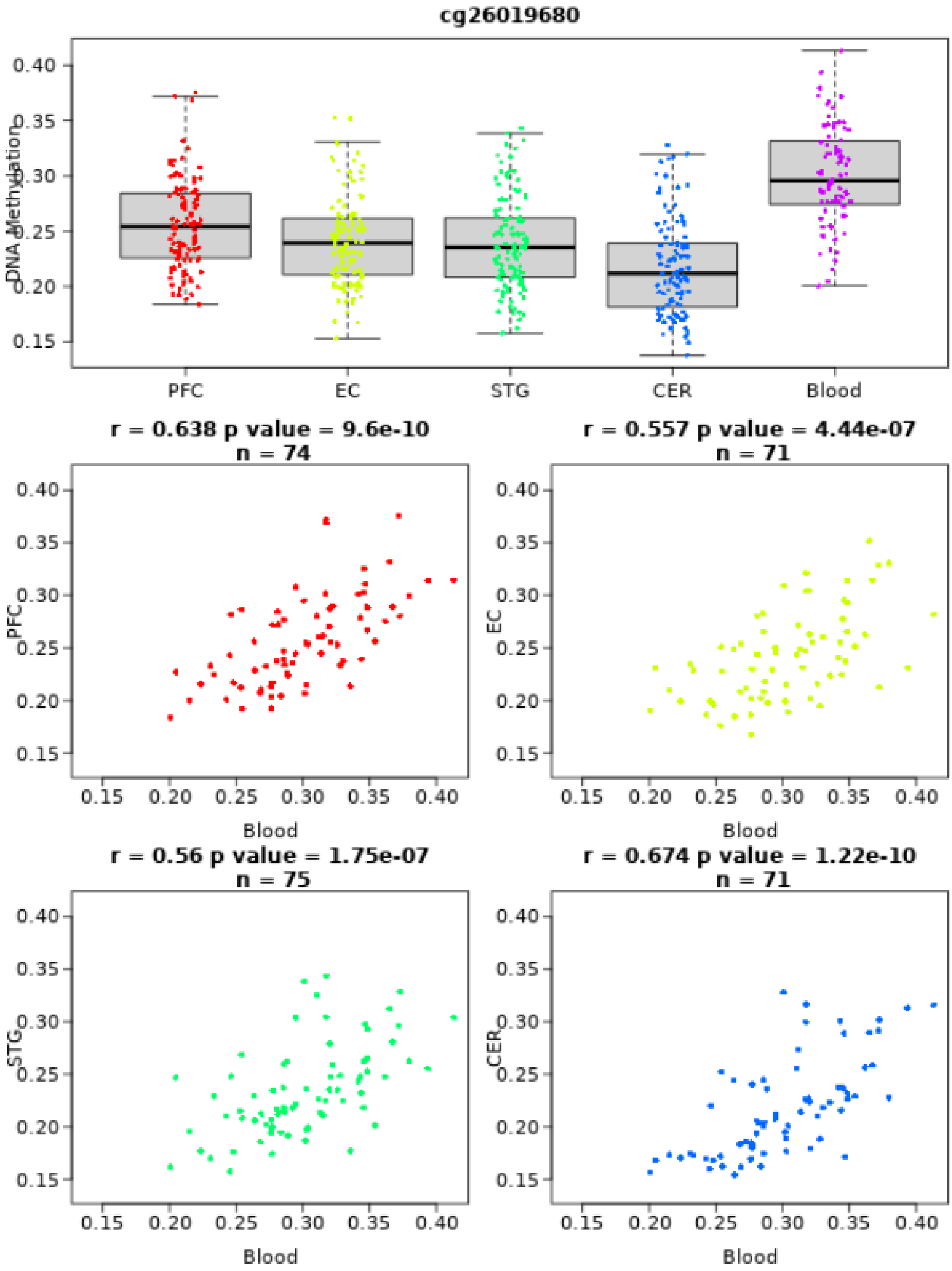


**Supplementary Figure 1** Brain-blood correlations for DNA methylation levels at cg26019680 located in promoter region of the PODXL2 gene. These figures were obtained using the Blood Brain DNA Methylation Comparison Tool (https://epigenetics.essex.ac.uk/bloodbrain/?probenameg=cg26019680). **Abbreviations** PFC: Prefrontal Cortex, EC: Entorhinal Cortex, STG: Superior Temporal Gyrus, CER: Cerebellum
